# Large-scale CSF proteome profiling identifies biomarkers for accurate diagnosis of Frontotemporal Dementia

**DOI:** 10.1101/2024.08.19.24312100

**Authors:** Yanaika S. Hok-A-Hin, Lisa Vermunt, Carel F.W. Peeters, Emma L. van der Ende, Sterre C.M. de Boer, Lieke H. Meeter, John C. van Swieten, William T. Hu, Alberto Lleó, Daniel Alcolea, Sebastiaan Engelborghs, Anne Sieben, Alice Chen-Plotkin, David J. Irwin, Wiesje M. van der Flier, Yolande A.L. Pijnenburg, Charlotte E. Teunissen, Marta del Campo

## Abstract

Diagnosis of Frontotemporal dementia (FTD) and the specific underlying neuropathologies (frontotemporal lobar degeneration; FTLD-Tau and FTLD-TDP) is challenging, and thus fluid biomarkers are needed to improve diagnostic accuracy. We used proximity extension assays to analyze 665 proteins in cerebrospinal fluid (CSF) samples from a multicenter cohort including patients with FTD (n = 189), Alzheimer’s Disease dementia (AD; n = 232), and cognitively unimpaired individuals (n = 196). In a subset, FTLD neuropathology was determined based on phenotype or genotype (FTLD-Tau = 87 and FTLD-TDP = 68). Forty three proteins were differentially regulated in FTD compared to controls and AD, reflecting axon development, regulation of synapse assembly, and cell-cell adhesion mediator activity pathways. Classification analysis identified a 14- and 13-CSF protein panel that discriminated FTD from controls (AUC: 0.96) or AD (AUC: 0.91). Custom multiplex panels confirmed the highly accurate discrimination between FTD and controls (AUCs > 0.96) or AD (AUCs > 0.88) in three validation cohorts, including one with autopsy confirmation (AUCs > 0.90). Six proteins were differentially regulated between FTLD-TDP and FTLD-Tau, but no reproducible classification model could be generated (AUC: 0.80). Overall, this study introduces novel FTD-specific biomarker panels with potential use in diagnostic setting.

## Introduction

Frontotemporal dementia (FTD) is the second most common form of young onset dementia (dementia with a symptom onset < 65 years old). FTD presents heterogeneously, comprising various clinical, neuropathological, and genetic forms. From a clinical perspective, FTD can present with either behavioral and social changes (behavioral variant FTD; bvFTD), language impairment (semantic variant primary progressive aphasia; svPPA and non-fluent variant PPA; nfvPPA), or motor dysfunction (corticobasal syndrome; CBS and progressive supranuclear palsy; PSP) [1–4]. FTD is caused by different underlying neuropathologies leading to frontal temporal lobar degeneration (FTLD). Approximately 50% of FTLD cases develop aggregates of the microtubule Tau protein (FTLD-Tau), while 45% is characterized by cytoplasmatic inclusion of the TDP-43 protein (FTLD-TDP). A small percentage of FTLD cases (5%) develop aggregates of the FUS protein (FTLD-FET) [5–7]. These pathologies can be predicted in familial forms of FTLD; where autosomal dominant mutations in the microtubule-associated protein tau (*MAPT)* gene lead to FTLD-Tau pathology, while mutations in the progranulin (*GRN)* or chromosome 9 open reading frame 72 (*C9orf72)* genes are associated with FTLD-TDP pathology [5, 8]. However, genetic FTD accounts only for 10-20% of FTLD individuals and in many families the mutation status is unknown and a new mutation arises [9, 10]. In sporadic cases, which are more common, the different neuropathological subtypes poorly correlate with the clinical presentation, with the exception of FTD accompanied with motor neuron disease and svPPA that shows a clinicopathological correlation with TDP-43 [11]. Thus, the different clinical phenotypes can have overlapping pathologies despite diverse genetic backgrounds, stressing the complexity of the disease and the importance of developing pathology-specific biomarkers, which is needed for targeted treatment.

Currently, there are no specific fluid biomarkers for the diagnosis of FTD, nor its neuropathological subtypes [12]. Given the overlapping clinical features with Alzheimer’s disease (AD) [13, 14], the analysis of the core AD-CSF biomarkers (i.e., amyloid-beta 1-40 and 1-42, Aβ1-40 and Aβ1-42 or the Aβ1-42/Aβ1-40 ratio; phosphorylated Tau181, pTau181; and total Tau, tTau) is often used to exclude AD diagnosis [15]. The concentrations of Neurofilament light (NfL) in CSF and blood are strongly increased in FTD but also in other neurodegenerative disorders as it is a biomarker reflecting neuroaxonal damage [16, 17]. Nevertheless, this biomarker is considered valuable for distinguishing FTD from non-neurodegenerative disorders, such as primary psychiatric diseases, and it also demonstrates good prognostic potential [18, 19]. Several studies have shown that the CSF pTau/tTau ratio could demarcate the main FTLD subtypes but with limited diagnostic accuracy [20, 21]. However, promising findings have been recently obtained with plasma GFAP/NfL ratio (FTLD-Tau vs. FTLD-TDP; AUC: 0.89), which requires additional validation in external cohorts [22]. Overall, there is an unmet need for fluid biomarkers specifically identifying FTD and its underlying pathologies, which is essential for accurate diagnosis, clinical trial inclusion, and to monitor the effects of treatments [23].

CSF proteomics offer the opportunity to study brain changes and associated processes *in vivo*. As extensively explored in AD, such analysis could offer insights into the disease’s etiology and reveal novel biomarker candidates [24–27]. Previous FTD studies using unbiased mass spectrometry (MS)-based technologies identified multiple biomarker candidates (e.g., YKL-40 in FTD and NPTXR in *GRN* mutation carriers) [28, 29]. However, subsequent validation efforts did not show sufficient diagnostic accuracy of these markers [30–33]. The clinical and pathological heterogeneity of FTD, switch in methodology between discovery and validation, and low sample sizes could explain the lack of validated biomarkers [28, 29]. Following a similar strategy to that previously applied for AD and DLB [27, 34], we here used the high-throughput immune-based proximity extension assay (PEA), to analyze the CSF proteome of an extensive and well-characterized cohort of patients with FTD presenting at memory clinics. We first aimed to identify CSF protein changes specifically associated with FTD and its main pathological subtypes (i.e., FTLD-Tau and FTLD-TDP). Next, we aimed to translate these findings into clinically feasible CSF protein panels to discriminate FTD and its pathological subtypes. Finally, we validated these panels in three independent cohorts, including one with autopsy confirmation.

## Results

### Subjects

An overview of the study is presented in Figure 1A. The discovery cohort included a total of 617 samples, all measured as part of previous CSF proteomic studies [27, 34]. Custom multiplex panels were developed and validated in two independent clinical cohorts (validation cohort 1: n = 161; validation cohort 2: n = 163) and one FTLD/AD autopsy-confirmed cohort (n = 100). The demographic characteristics for all cohorts are described in Table 1. Controls were younger compared to other diagnostic groups in all cohorts, except in the FTLD/AD autopsy cohort. Furthermore, patients with AD were older, and patients with FTLD were overall younger in the FTLD/AD autopsy cohort compared to the other cohorts.

**Figure 1.**
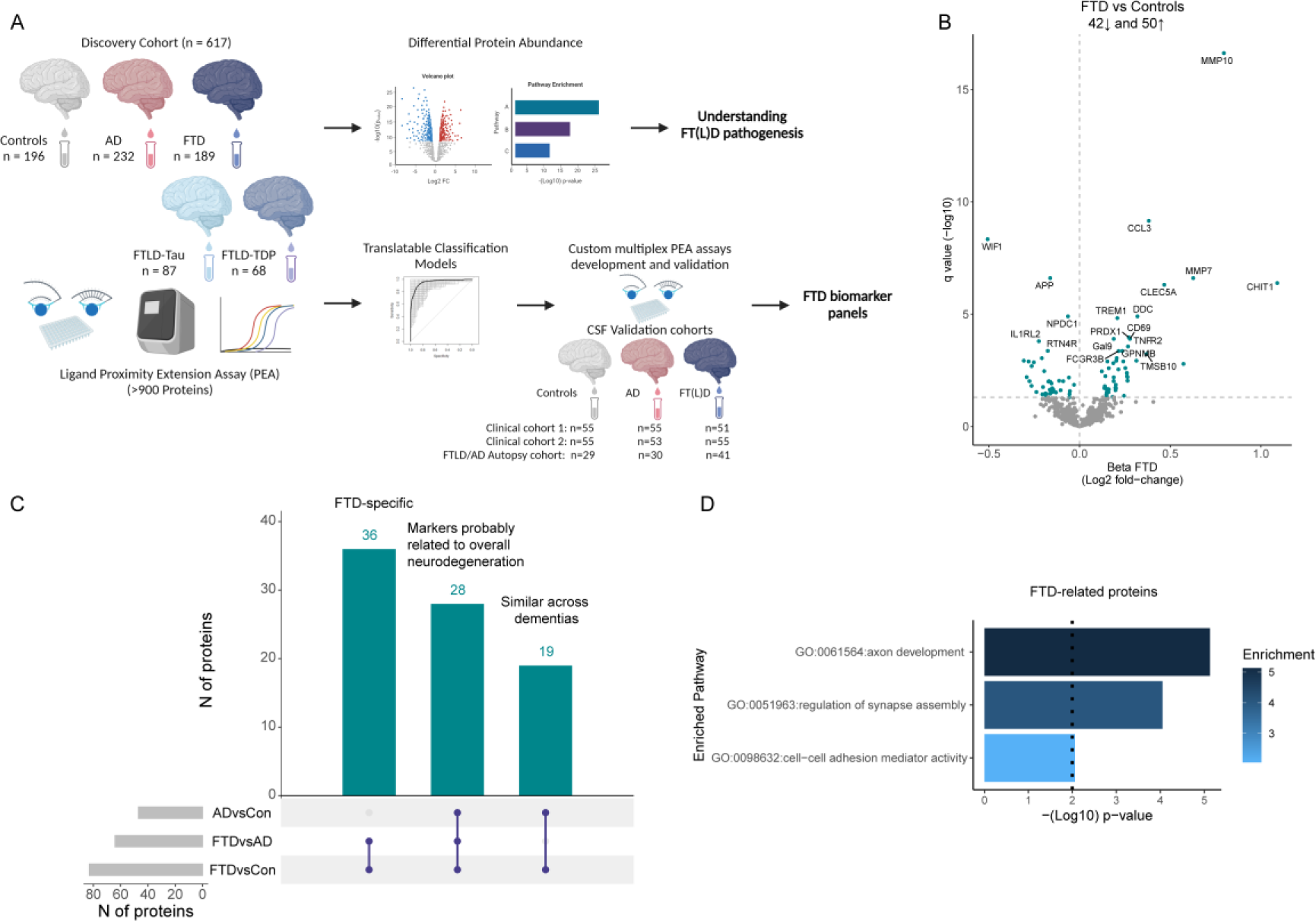
Overview study design and differential abundance of CSF proteins in FTD. A) More than 900 proteins were measured using the antibody-based PEA technology in CSF from 196 cognitively unimpaired controls (white), 189 FTD (blue), and 232 AD (red) patients. Differential protein abundance was investigated and classification models were constructed. Custom PEA assays using the proteins identified in the classification models were developed and validated in three independent cohorts, including an FTLD/AD autopsy cohort. B) Volcano plots show that 93 CSF proteins were differentially regulated between FTD and controls. Each dot represents a protein. The beta coefficients (log2 fold-change) are plotted versus q values (-log10-transformed). Proteins significantly dysregulated after adjusting for false discovery rate (FDR, q < 0.05) are depicted in blue. The total number of proteins that are down-regulated (n = 42, left) or up-regulated (n = 51, right) are depicted. Horizontal dotted line indicate the significance threshold. C) UpSet plot shows proteins dysregulated between FTD and controls and also dysregulated between FTD and AD or AD and controls. D) Bar graphs depicting the biological pathways enriched in protein specifically dysregulated in FTD. Dotted line represents the significant threshold (p < 0.01). The corresponding GO number and biological process is depicted on the left side. Stronger colors represent higher significant enrichment.

**Table 1.** Cohort characteristics.

|  | Discovery Cohort |  |  | Validation Cohort 1 |  |  | Validation Cohort 2 |  |  | FTLD/AD Autopsy Cohort |  |  |
| --- | --- | --- | --- | --- | --- | --- | --- | --- | --- | --- | --- | --- |
|  | CON | FTD | AD | CON | FTD | AD | CON | FTD | AD | CON | aFTLD | aAD |
| n | 196 | 189 | 232 | 55 | 51 | 55 | 55 | 55 | 53 | 29 | 41 | 30 |
| Sex, Female n (%) | 72 (36.7) | 93 (49) | 96 (41) | 22 (40) | 17 (33) | 23 (42) | 32 (58) | 21 (38) | 34 (64) | 13 (45) | 20 (49) | 15 (50) |
| Age | 58 (53 - 63)<sup>a,b</sup> | 65 (59 - 70)<sup>c</sup> | 66 (59 - 72)<sup>c</sup> | 58 (56 - 62)<sup>a,b</sup> | 63 (59 - 69)<sup>c</sup> | 66 (62 - 70)<sup>c</sup> | 61 (58 - 67)<sup>a,b</sup> | 76 (71 - 79)<sup>c</sup> | 74 (68 - 76)<sup>c</sup> | 64 (61 - 66) | 64 (55 - 71)<sup>b</sup> | 72 (64 - 76)<sup>a,c</sup> |
| MMSE | 28 (27 - 29)<sup>a,b</sup> | 25 (22 - 27)<sup>b,c</sup> | 21 (17 - 24)<sup>a,c</sup> | 29 (27 - 30)<sup>a,b</sup> | 25 (23 - 27)<sup>b,c</sup> | 20 (18 - 23)<sup>a,c</sup> | 29 (29 - 30)<sup>a,b</sup> | 21 (18 - 25)<sup>c</sup> | 22 (19 - 25)<sup>c</sup> | 29.5 (29 - 30) | 20 (14 - 26) | 14 (7 - 21) |
| CSF AD biomarker +, n (%) | 0 (0) | 35 (19) | 230 (99) | 3 (5.5) | 9 (17.6) | 49 (89) | 0 (0) | 1 (2) | 50 (194.3) | 1 (3) | 19 (46) | 29 (97) |
| APOE4 carrier, n (%) | 48 (25) | 46 (24) | 132 (57) | 15 (27) | 13 (26) | 39 (71) | 14 (26) | 8 (15) | 22 (41.5) |  |  |  |
| FTLD-Subtypes, n |  | 87 FTLD-Tau<br>67 FTLD-TDP |  |  |  |  |  |  |  |  | 7 aFTLD-Tau<br>31 aFTLD-TDP<br>3 aFTLD-UPS |  |

Continuous variables are presented as median ± interquartile range and dichotomous data as the number of cases with a percentage of the total (%). Differences between groups were determined using the Kruskal Wallis test with Bonferroni correction or Chi-squared test.
a, $p < 0.05$ compared to FT(L)D
b, $p < 0.05$ compared to AD
c, $p < 0.05$ compared to CON
Abbreviations: CON = Controls, FTD = frontotemporal dementia, AD = Alzheimer's disease, MMSE = mini-mental state examination, CSF = cerebrospinal fluid, FTLD = frontotemporal lobar degeneration, TDP = Transactive response DNA binding protein of 43, FUS = fused in sarcoma, aFTLD = autopsy confirmed FTLD, aAD = autopsy confirmed AD.

### CSF proteins are differentially regulated in FTD compared to controls and AD

CSF proteome profiling revealed 92 proteins differentially regulated between FTD and controls, of which 42 proteins were decreased and 50 proteins were increased (*q* < 0.05; Figure 1B, Supplementary Table 2). The top 5 differentially regulated proteins related to FTD (median *q*: 2.5^-07^) are involved in the regulation of the Wnt signaling pathway (WIF1), tissue remodeling (MMP7), neuronal biosynthesis (APP), cell proliferation (NPDC1), and innate immunity (IL1RL2). Next, we sought to determine whether the protein changes identified were especially related to FTD. We show that 36 proteins (40%) were uniquely dysregulated in FTD compared to controls and AD (e.g., WIF1, ROBO2, and SLITRK2; Figure 1C). Nineteen proteins (20%) were dysregulated in controls compared to FTD and AD, which likely represent general dementia processes (e.g., CHIT1, MMP10, and MMP12; Figure 1C). Twenty-eight proteins (30%) were dysregulated across all comparisons: controls versus FTD and AD, and FTD versus AD. Most of these proteins (21 out of 28; e.g., MIF, ITGB2, and sTREM1) were to some extent increased in FTD but were more prominently dysregulated in AD, indicating that they are likely more related to AD pathophysiological processes. However, 7 out of the 28 proteins (e.g., CHL1, GPC1, CNTN5, HBEGF, CLSTN2, GCP5) showed an opposite direction of change in FTD and AD and are likely differently involved in the pathophysiology of these types of dementias. Functional enrichment analysis of the proteins related to FTD pathophysiology (i.e., those that were increased in FTD as well as those that were specifically dysregulated in FTD compared to AD, n = 43) revealed associations with axon development, regulation of synapse assembly, and cell-cell adhesion mediator activity (Figure 1D).

### CSF proteins are differentially regulated in FTLD-Tau or FTLD-TDP subtypes

Next, we investigated which proteins dysregulations charachterized the FTLD-Tau and FTLD-TDP subtypes. Compared to controls, we observed 60 proteins dysregulated in FTLD-Tau (top 5: MMP10, DDC, CCL3, MMP7, and WIF1; median *q* 4.2^-08^, q < 0.05; Figure 2A, Supplementary Table 3) and 120 proteins that were dysregulated in FTLD-TDP (top 5: APP, NPDC1, WIF1, B4GAT1, and ROBO2; median *q 1.2^-06^*; Figure 2B, Supplementary Table 4). When comparing FTLD-Tau to FTLD-TDP, we observed that 6 proteins (COCH, Siglec9, VSIG4, GRN, CD84, and C1QTNF1) were dysregulated between these groups (*q* < 0.05; Figure 2C, Supplementary Table 5). This number increased to 185 dysregulated proteins when nominal significance was considered (i.e., p < 0.05, Supplementary Table 5). To show which proteins are specifically associated with one of the FTLD subtypes, we visualized the outcomes of three comparisons in an upset plot (Figure 2D). Here, we identified three proteins uniquely related to FTLD-Tau (i.e., all increased in FTLD-Tau compared to controls and FTLD-TDP; Figure 2E). These proteins play a role in various processes, including phagocytosis and the NLRP3 inflammasome (VSIG4), cell adhesion processes (Siglec9), and involvement in both the innate and adaptive immune responses (CD84)[35–37]. Two proteins were uniquely related to FTLD-TDP (i.e., both decreased in FTLD-TDP compared to controls and FTLD-Tau; Figure 2E), which are related to lysosomal functioning (GRN) and the immune response (C1QTNF1)[38, 39]. Interestingly, one protein was uniquely dysregulated between FTLD-Tau and FTLD-TDP (COCH; Figure 2E), with higher levels in FTLD-Tau. This protein plays a role in the modulation of cell shapes [40].

**Figure 2.**
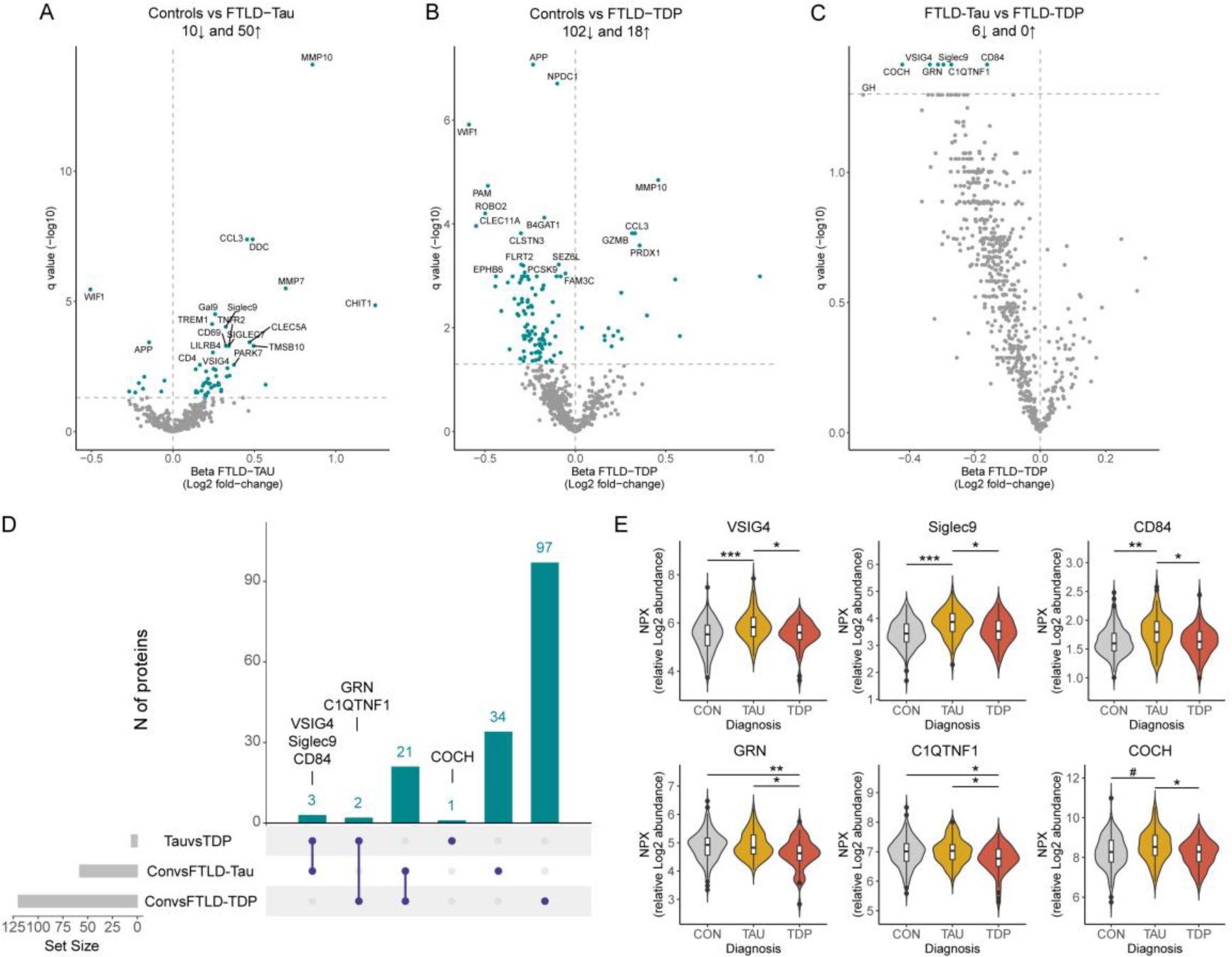
Differential abundance of CSF proteins in FTLD subtypes. Volcano plots show CSF proteins differentially regulated between patients with FTLD-Tau (n = 87; A) or FTLD-TDP (n = 68; B) and controls (n = 196) and between these neuropathological subtypes (C). Each dot represents a protein. The beta coefficients (log2 fold-change) are plotted versus q values (-log10- transformed). Proteins significantly dysregulated after adjusting for false discovery rate (FDR, q < 0.05) are depicted in blue. The total number of proteins that are down-regulated (left) or up-regulated (right) is indicated. D) UpSet plot depicts proteins dysregulated between FTLD-Tau, FTLD-TDP, and controls groups. E) Violins represent the abundance (log2 NPX) of the CSF proteins that were uniquely dysregulated in FTLD-Tau and FTLD-TDP or between the subtypes. Boxplot within the violin indicates the median and interquartile range of the protein abundance. # p < 0.05, *q < 0.05, **q < 0.01, ***q < 0.001. Abbreviations: CON, cognitively unimpaired controls; TDP, Transactive response DNA binding protein of 43.

### CSF protein panels discriminate with high accuracy FTD from controls and AD

Next, we aimed to identify CSF biomarkers specific for FTD. One of the strongest proteins specifically dysregulated in FTD was WIF1, however, its performance as single marker to discriminate FTD from controls was moderate (AUC: 0.794, 95% CI: 0.75-0.84). Thus, we next performed a classification analysis, followed by internal cross-validation, to investigate whether a combination of proteins could discriminate FTD from controls with higher accuracy (CSF panels, Figure 1A). We identified a panel of 14 CSF proteins that discriminated FTD from controls with high accuracy (FTD diagnostic panel; AUC: 0.96, 95% CI: 0.91-0.99; Figure 3A), which was comparable to the performance of CSF NfL (AUC: 0.96, 95% CI: 0.92-1; Figure 3B). The model included proteins that were dysregulated specifically in FTD (i.e., WIF1, MMP7, GAL, VEGFA, NPDC1, APP, and CCL11; Supplementary Figure 1), as wel as proteins related to common neurodegenerative processes and other neurodegenerative dementias (i.e., MMP1, MMP10, CHIT1, CCL3, PRDX1, and DDC; Supplementary Figure 1), or reported to be associated with AD (i.e., CLEC5A; Supplementary Figure 1). The performance of the FTD diagnostic panel to discriminate FTD from AD was considerably lower (FTD diagnostic panel AUC: 0.77, 95% CI: 0.67-0.86; for comparison: CSF NFL AUC: 0.80, 95% CI: 0.72-0.88, Figure 3B). We thus next investigated whether we could find a combination of proteins that optimally discriminated FTD from AD. A panel consisting of 13 CSF proteins was identified that discriminated FTD from AD with high accuracy (FTD differential diagnostic panel; AUC: 0.91, 95% CI: 0.85-0.96; Figure 3C). This panel contained proteins associated with FTD (i.e., GZMB, MMP7, CCL11, NPDC1, PLTP, and APEX1; Supplementary Figure 2) but also proteins associated with AD (i.e., ABL1, ENO2, ITGB2, SMOC2, and THOP1; Supplementary Figure 2), as also reported in our previous AD focused PEA study [27]. The model also included two proteins with no differences across groups, likely due to the model’s adjustment for inter-individual variability (i.e., VEGFR3 and KAZALD1; Supplementary Figure 2) [41]. Of note, three proteins (i.e., MMP7, NPDC1, and CCL11) were included in both the FTD diagnostic and FTD differential diagnostic panel. The FTD-related proteins from these panels are associated with diverse biological pathways including inflammatory processes (GZMB), tissue remodeling (MMP1, MMP7), cell proliferation (NPDC1), oxidative stress (APEX1), vascular functioning (VEGFA), and neuronal biosynthesis and functioning (APP and GAL)[42–46].

**Figure 3.**
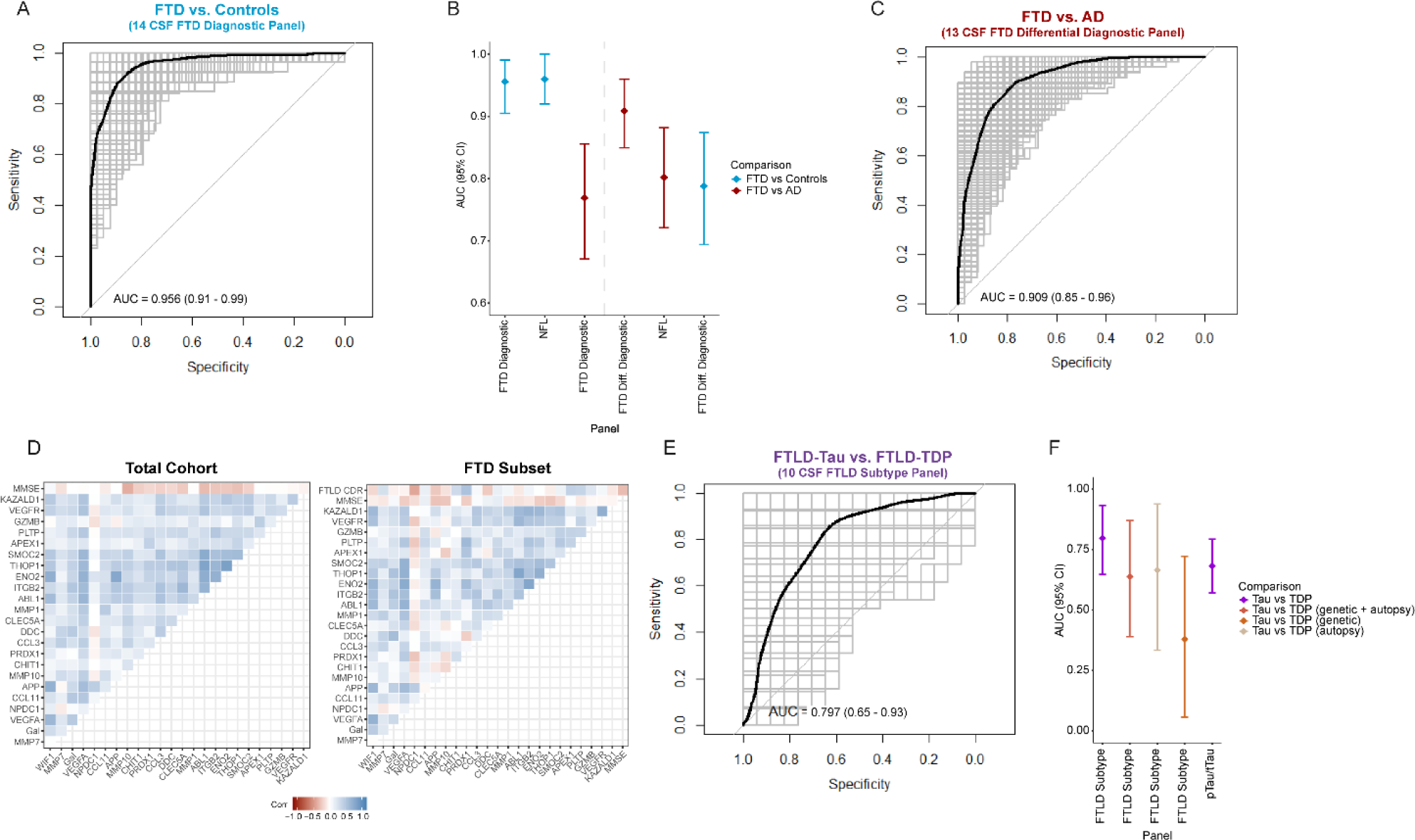
CSF biomarker panels for specific diagnosis of FTD. A) Receiver operating characteristic (ROC) curves depict the performance of 14 CSF biomarker panel discriminating FTD from controls. Black line is the mean area under the curve (AUC) overall re-samplings (1000 repeats of 5-fold cross-validation, grey lines). B) Forest plot shows the different AUC and 95% confidence interval for the 14 and 13 CSF biomarker panels, and NFL to discriminate between FTD and controls (blue) or AD (red). C) Correlation matrix heatmap representing the Spearman’s correlation coefficient in-between the proteins selected in each panel, MMSE score, and FTLD-CDR scores in the total cohorts, and in a subset of the FTD group (n = 62). ROC curves depict the performance of 13 CSF biomarker panel discriminating FTD from AD (D) and FTLD-Tau from FTLD-TDP (E). Forest plot shows the different AUC and 95% confidence interval for the 10 CSF biomarker panels, and pTau/tTau ratio to discriminate between neuropathological subtypes (purple), subtypes that are genetic and/or autopsy confirmed (brown), subtypes which are genetic confirmed (coral), and subtypes which are autopsy confirmed (beige).

Next, we sought to determine if the proteins included in our FTD panels are associated with disease severity and cognition as measured by FTLD-CDR and MMSE scores. In the FTD patients, we observed that PRDX1 (*Rho* = 0.47, p < 0.001) had a moderate to strong correlation while NPDC1 (*Rho* = -0.37, p < 0.001), APEX (*Rho* = 0.35, p < 0.01), PLTP (*Rho* = 0.33, p < 0.01), and CHIT1 (*Rho* = 0.28, p < 0.05) showed a moderate correlation with FTLD-CDR scores (Figure 3D). In the total cohort, we show that the proteins associated with AD (ABL1, ENO2, ITGB2, SMOC2, and THOP1) together with MMP10 had the strongest correlations with MMSE scores (*Rho’s* between -0.19 and -0.34, all p < 0.001; Figure 3D).

### CSF protein panel to discriminate the FTLD subtypes

We identified a panel of 10 CSF proteins that discriminated between FTLD-Tau and FTLD-TDP with moderate performance (AUC: 0.797, 95% CI: 0.65-0.93; Figure 3E). Despite the performance being superior to the pTau/tTau ratio (AUC: 0.68, 95% CI: 0.57-0.79; Figure 3F), the large CI reflects high variability in the cross-validation procedures and thus poor robustness. This model included proteins that were specifically dysregulated in FTLD-Tau (i.e., VSIG4, CD84, and Siglec9), or FTLD-TDP (i.e., GRN) and those that were nominally or significantly changed between subtypes (i.e., COCH, GH, CST5, and PRSS8; Supplementary Fig 3), as well as proteins that were similar across groups (i.e., IFNLR1 and ANGPT2; Supplementary Fig 3).The performance of this model did not improve after stratifying the FTLD groups for genetic and sporadic cases (AUCs: 0.37 – 0.64; Figure 3F).

### Validation of FTD Panels using custom PEA assays in three independent cohorts

We next developed custom multiplex-PEA assays containing 18 of the 24 proteins from the FTD panels described above (i.e. FTD diagnostic panel: 9 proteins; FTD differential diagnostic panel: 12 proteins; 3 proteins present in both panels). Six proteins could not be included due to technical limitations. The custom PEA assays (custom panels) showed over 90% detectability of the proteins in patient samples, with low coefficients of variation (CVs; <5% intra-CV and <3% inter-CV; Supplementary Table 1). Next, we validated these custom panels in three independent cohorts including one with autopsy confirmation. The proteińs fold changes obtained in the comparisons between FTD and controls significantly correlated between those detected in the discovery and two validation cohorts (clinical cohort 1: *Rho* = 0.45, p > 0.05, clinical cohort 2: *Rho* = 0.65, p < 0.05 and FTLD/AD autopsy cohort: *Rho* = 0.93, p < 0.001; Supplementary Figure 4A). In addition, the fold changes of WIF1 and MMP10 were strong and replicated across all cohorts (Figure 4A). Similarly, strong correlations of the protein fold changes for the difference between FTD and AD patients were observed between the discovery cohort and all three validation cohorts (clinical cohort 1: *Rho* = 0.69, p < 0.05; clinical cohort 2: *Rho* = 0.78, p < 0.01; FTLD/AD autopsy cohort: *Rho* = 0.87, p < 0.001, Supplementary Figure 4B). Noteworthy, we observed that the fold changes of some proteins lost significance after correction for multiple testing in clinical cohort 1 and clinical cohort 2, whereas several proteins showed their strongest effects in the FTLD/AD autopsy cohort (e.g., THOP1, MMP7, ENO2, CCL11, and KAZALD1; Figure 4A).

**Figure 4.**
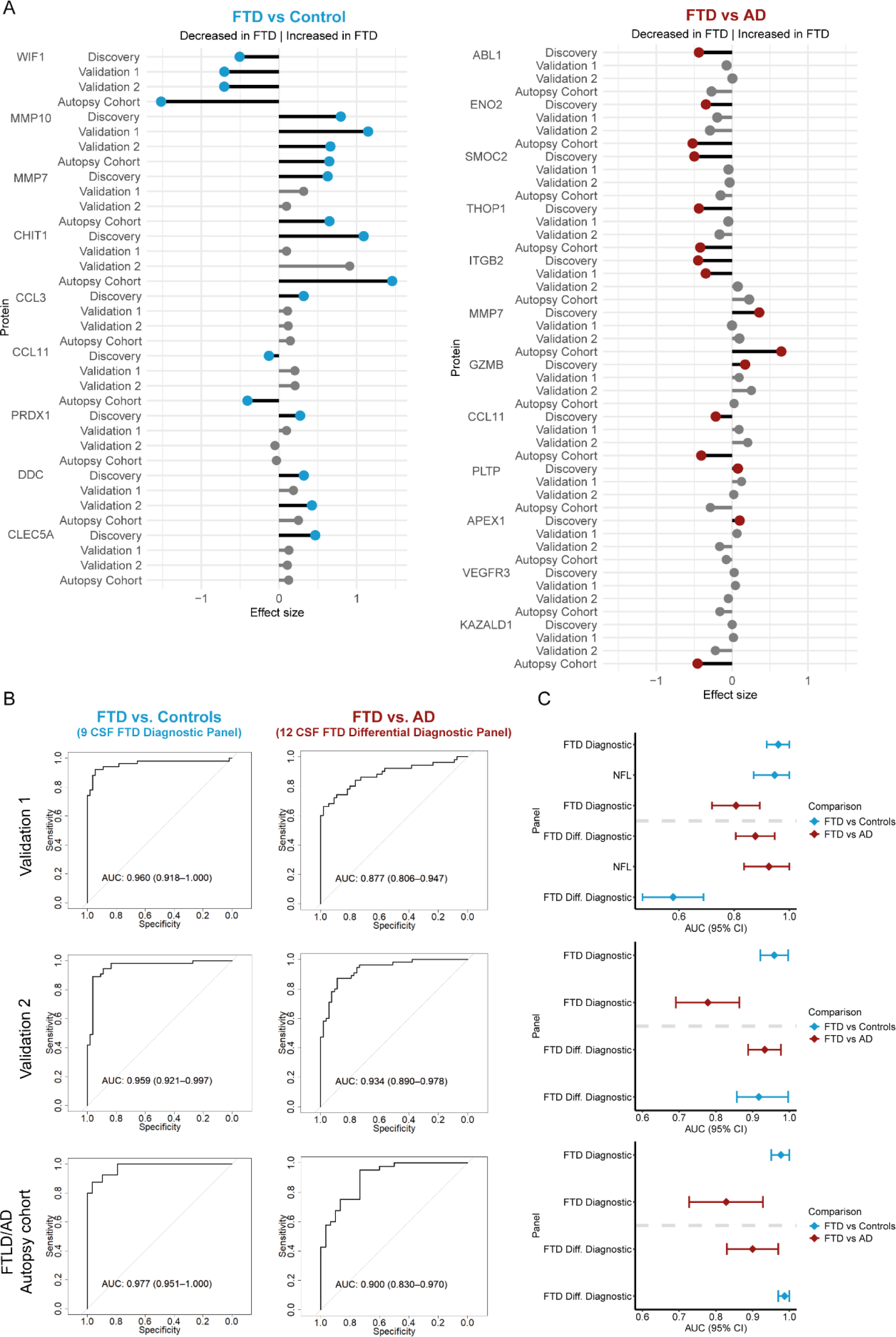
Development and validation of custom CSF biomarker panels for FTD diagnosis in independent cohorts. A) Lollipopplots depict the beta-coefficients obtained in the discovery phase in parallel to the beta-coefficients of the custom assays in clinical validation cohorts 1 and 2 and the autopsy cohort. Grey dots shows proteins that did not remain significant after correction for multiple testing. B) Receiver operating characteristic (ROC) curves showing the performance of the CSF biomarker panel discriminating FTD from controls or AD using the custom assays across the two clinical and one autopsy validation cohort. Inserts outline corresponding AUC and 95% CI. C) Forest plots depict the different AUC and 95% CI obtained with the CSF FTD biomarker panels or CSF NfL in the comparison between FTD and controls (blue) or AD (red).

The FTD diagnostic panel (containing 9 out of the 14 proteins) confirmed discrimination of FTD from controls with high accuracy in all validation cohorts (clinical cohort 1: AUC = 0.96, 95% CI: 0.92-1; clinical cohort 2: AUC = 0.96, 95% CI: 0.92-0.99; FTLD/AD autopsy cohort: AUC = 0.98, 95% CI: 0.95-1; Figure 4B) and was similar to CSF NfL (clinical cohort 1: AUC = 0.95, 95% CI: 0.87-1; Figure 4C). In addition, the FTD differential diagnostic panel (containing 12 out of the 13 proteins) could again discriminate FTD from AD with high accuracy in all three validation cohorts (clinical cohort 1: AUC = 0.88, 95% CI: 0.81-0.95; clinical cohort 2: AUC = 0.93, 95% CI: 0.89-0.98; FTLD/AD autopsy cohort: AUC = 0.90, 95% CI: 0.83-0.97; Figure 4B), which was similar to NfL (clinical cohort 1, AUC = 0.93, 95% CI: 0.84-1; Figure 4C). Thus, we replicated the discriminative performance as we observed in the discovery cohort, both for the FTD diagnostic and differential diagnostic panel.

## Discussion

This study identified novel and specific protein changes for FTD and its main pathological subtypes. These findings were translated into two CSF biomarker panels discriminating FTD from controls and AD patients with high accuracies (AUCs > 0.9). These results were validated by clinically feasible panels that measured the selected proteins in three independent cohorts, including one with autopsy confirmation. We identified proteins specifically associated with FTLD-Tau or FTLD-TDP groups, but we could not identify a marker or combination of markers to robustly discriminate between these FTLD subgroups. The FTD proteins and classification panels identified in this study reflect a broad range of different biological processes associated to FTD and its biological subtypes including, inflammatory processes, regulation of synapsis, lysosomal functioning, tissue remodeling, and oxidative stress.

Fluid biomarkers specifically associated with FT(L)D pathophysiology are needed to improve diagnostic accuracy, for clinical trial inclusion, and to monitor treatment effects [12]. We have performed a large multicenter FTD proteomics study, including patients with different FTLD neuropathologies as well as a group with AD dementia. Up to 92 proteins were dysregulated between FTD and controls. By comparing the proteins profiles to AD patients, we observed that 40% of these proteins were uniquely dysregulated in FTD (e.g., WIF1, ROBO2, and SLITRK2). Some proteins have been associated with FTD in previous CSF antibody-based proteomics (VEGFA [47]), or FTLD brain proteomics studies (ADAM23 [48–50] and WIF1 [51]). We also identified a subset of CSF proteins (30%) that were differentially regulated in both patients with FTD and AD compared to controls but with a different protein abundance between FTD and AD. Despite most of these being more prominently dysregulated in AD (e.g., SDC4, ITGB2, MIF, and sTREM1)[27, 52, 53], a small subset of proteins showed an opposite effect between these dementias (e.g., CHL1, GPC1, and CNTN5). Previous studies additionally detected decreased levels of CHL1 in FTD and PSP using alternative platforms, which further supports the validity of our findings [54, 55]. In addition, we detected a subset of proteins (20%) likely reflecting overall neurodegenerative process as they were similarly dysregulated in AD and FTD compared with controls. For instance, CHIT1 and MMP10 protein levels were increased in FTD and AD, as reported previously [27, 47, 56–61]. This study identifies numerous CSF proteins dysregulated in FTD. However, due to the inclusion of an AD group, we uncovered that half (43 out of 92) of the proteins are specifically associated with FTD, highlighting the importance of comparing to both controls and similar neurological diseases in biomarker studies. Among the proteins specifically associated with FTD, the ones showing the strongest effects were: WIF1, MMP7, APP, NPDC1, and IL1RL2. WIF1 is an inhibitory protein involved in the Wnt signaling cascade, a dysregulation of this pathway has been implicated in tau phosphorylation and other neuronal processes (e.g., neurogenesis, synaptic health and plasticity) [62, 63]. In addition, WIF1 was dysregulated in FTLD frontal cortex tissue [51], supporting its role within FTD pathophysiology, however, its specific function in relation to FTD remains to be elucidated. Enrichment analysis showed that proteins specifically dysregulated in FTD were enriched in biological pathways associated to axon development, regulation of synapse assembly, and cell-cell adhesion mediator activity. This is in line with previous unbiased brain and CSF proteomics studies, and multiplex CSF analysis showing a dysregulation of similar pathways in FTLD [29, 48, 49, 64, 65].

We additionally investigated the CSF proteome in the main FTLD pathological subtypes. Despite that many CSF proteins were dysregulated in FTLD-Tau (60 proteins) or FTLD-TDP (120 proteins) compared to controls, overlapping analysis across all comparisons revealed three proteins related to FTLD-Tau (VSIG4, Siglec9, and CD84), two proteins related to FTLD-TDP (GRN and C1QTNF1), and one protein uniquely changed between the neuropathological subtypes (COCH). Given the established association between *GRN* mutations and TDP pathology, the decrease of GRN (progranulin) protein levels observed in FTLD-TDP aligns with previous GRN reductions in CSF and plasma of FTD-*GRN* mutation carriers [66, 67]. It should be noted that the FTLD-TDP group included a low number of *GRN* mutation carriers (11%). Considering that we observed decreased GRN levels in the total FTLD-TDP group, these results highlight that lysosomal dysfunction is an important feature in TDP pathology, which is additionally supported by post-mortem analysis [68, 69]. Furthermore, we also uncoverd novel markers such as VSIG4. This protein is a phagocytic receptor involved in various inflammatory responses and can mediate the NLRP3 inflammasome activation [35]. Since the NLRP3 inflammasome plays a role within tau pathophysiology, this protein seems promising to reflect ongoing activation of the NLRP3 inflammasome in FTD cases [70]. To the best of our knowledge, many of the subtype-specific proteins identified in this study are novel and have not yet been linked to the main FTD pathologies or genetic subtypes in previous proteomics studies [28, 29, 48, 49]. Thus, these proteins require further investigation in CSF and brain tissue, to examine their relevance to ongoing tau or TDP43 pathophysiologies.

To translate the CSF proteome findings into practical biomarker tools for routine diagnostics or clinical trials, we applied classification analyses and identified two panels of 14 and 13 CSF proteins that can discriminate FTD from controls and AD dementia with high accuracies (AUCs of 0.96 and 0.91, respectively). Thereafter, custom multiplex assays were developed and validated in three independent cohorts. The fold changes for each protein correlated well between discovery and validation cohorts (*Rho’s* between 0.45-0.93). The high discriminative values were confirmed in all three cohorts for both panels (AUCs > 0.877), supporting the robustness of our findings. The relevance of these FTD panels is additionally supported by the association of some markers (e.g., MMP10, PLTP, PRDX1, and NPDC1) with clinical parameters such as cognitive functioning or disease severity of FTD. Noteworthy, our novel FTD panels have similar discriminative performance compared to NfL. However, while NfL reflects neuroaxonal damage, our CSF panels offer a more comprehensive depiction of the underlying biological processes in FTD. Furthermore, within our FTD panels, we detected positive correlations of APEX, PRDX1, and CHIT1 with FTLD-CDR scores. These proteins are related to oxidative stress and reactive microglial, processes that underly FTD pathogenesis, and thus could be relevant for disease staging [71, 72]. In addition, in agreement with previous studies, we observed that increased MMP10 concentrations were associated with worse cognitive performance [27, 56]. This could potentially be explained by the expression of MMP10 by microglial cells in the brain, as elevated MMP10 levels contribute to ongoing inflammatory responses leading to axonal damage, potentially leading to functional deficits such as cognitive impairment [73]. Overall, these associations provide further support for the potential value of these panels in clinical settings and trial contexts.

Lastly, we identified a classification signature of 10 CSF proteins to discriminate the two main FTLD subtypes (AUC: 0.8), with higher performance than the pTau/tTau ratio (AUC: 0.68). However, the moderate performance together with the large confidence intervals suggest that the model is rather unstable and may vary depending on the samples included. This variation could be explained bij the inclusion of different FTD biochemical profiles, which likely differ between sporadic and familial cases with the same proteopathy, or even between genetic backgrounds of the same pathological subtypes (e.g., GRN and C9orf72) [12, 54, 74]. Despite the unprecedented number of FTLD samples and proteins analyzed in this study, we detected few markers associated to specific pathological subtypes. However, many proteins showed nominal significant differences, underscoring the need for larger sample sizes to identify additional FTLD-specific biomarkers. This also highlights the biological heterogeneity within each FTLD subtype and the need to analyze well-characterized FTD cohorts with homogeneous groups, calling for collaborative studies across different centers [12].

This study is not without limitations. The targeted proteomic approach employed in this study misses relevant proteins that might have been measured using unbiased MS methods (e.g., GFAP and neuropentraxins). However, our study still covered > 600 proteins covering a wide range of biological mechanisms and the workflow employed allowed us to swiftly translate our proteomic discovery findings into custom immunoassays for subsequent validation. Noteworthy, potential FTD misdiagnosis may influence biomarker results. However, diagnoses were made in specialized memory clinics and 47% of FTD individuals were either pathologically or genetically confirmed, and results were further validated in three independent cohorts. Still, it would be of interest to evaluate the performance of these novel FTD panels in other disorders sharing clinical and/or neuropathological features with FTD, such as primary psychiatric disorder or amyotrophic lateral sclerosis.

## Conclusion

This study identified CSF proteome changes specifically associated to FTD. We have translated these findings into CSF protein panels that can accurately discriminate FTD from controls and AD in multiple cohorts. Furthermore, we detect protein changes specifically associated to FTLD-Tau or FTD-TDP, although larger and more homogeneous cohorts will have more power to discriminate between these main pathological subtypes. The panels developed within this study could prove valuable for diagnosis or to monitor effects of treatments in clinical trials. The antibody-based technology employed in this study allowed us to efficiently translate our discovery findings into custom multiplex panels for further clinical validation showing reproducible findings. This workflow applied here could also be helpful for the development of fluid biomarkers in other human matrices (e.g., blood) and other biomedical fields beyond neurodegenerative dementias.

## Method

### Participants

As part of our previous work [27], the discovery cohort (total = 617; Table 1) included CSF samples from patients diagnosed with FTD (n = 189), AD dementia (n = 232), and cognitively normal individuals (CON, n = 196, Table 1). Most of the samples were selected from the Amsterdam Dementia Cohort (ADC; 120 FTD, 214 AD, 190 CON)[75]. To enrich for samples from patients with confirmed FTD, additional cases from the Center for Neurodegenerative Disease Research at the University of Pennsylvania (Penn; 46 FTD, 18 AD, 6 CON), Erasmus Medical Center (19 FTD), and the Goizueta Alzheimer’s Disease Research Center at Emory University (4 FTD) were selected. For a subset of the FTD patients (82%), the underlying neuropathology was known or could be predicted based on specific clinical diagnosis (FTLD-Tau = 87, FTLD-TDP = 67). FTLD-Tau was confirmed based on autopsy (n = 17), *MAPT* mutation (n = 16), and further enriched with patients clinically diagnosed with PSP (n = 31) and CBS (n = 23), which primarily associates with Tau neuropathology [76]. The FTLD-TDP group included autopsy-confirmed cases (n = 25) and patients with *GRN* (n = 8) or *C9orf72* (n = 23) mutations, and further enriched with patients clinically diagnosed with svPPA (n = 11), which have a high likelihood of having TDP pathology [11]. Three additional validation cohorts were included for validation of the custom panels: two clinical cohorts, from the ADC (51 FTD, 55 AD, and 55 controls) and Sant Pau Initiative on Neurodegeneration (SPIN; 55 FTD, 53 AD, and 55 controls)[77] and one FTLD/AD autopsy confirmed cohort from BIODEM, U Antwerp and the neurobiobank of the Institute Born-Bunge (IBB) / UAntwerp (41 autopsy-FTLD; aFTLD, including 7 aFTLD-Tau and 31 aFTLD-TDP, 3 aFTLD-UPS, and 30 autopsy-AD; aAD). Additionally, 29 cognitively unimpaired controls from BIODEM-UAntwerp were included in this cohort but these were not autopsy-confirmed.

All participants underwent standard neurological screening and cognitive testing. FTD and AD diagnoses were made according to international consensus criteria [1–4, 78]. The autopsy-confirmed cohort included cases with a definite diagnosis according to international neuropathological examination guidelines for FTLD [79], and AD [80]. Mini-mental state examination (MMSE) was used as a measurement for global cognition in all groups. In addition, the FTLD CDR® plus NACC score, a clinical measure specific for FTD disease severity, for a subset of FTD cases from the ADC (discovery cohort, n = 62; clinical cohort 1, n = 44) [81]. The control group included individuals with subjective cognitive decline, which scored normally on cognitive examinations with negative AD CSF biomarkers profile.

CSF was collected and biobanked according to established protocols [82]. Concentrations of CSF Aβ42, pTau, and tTau were used to support AD diagnosis. These were analyzed locally as part of the diagnostic work-up using commercially available kits (ADC, Erasmus MC, and IBB: ELISA Innotest Aβ(1-42), hTAUAg, pTau (181P; Fujirebio, Ghent), or ADC: Elecsys Aβ42, t-tau and p-tau (181P) CSF assays (Roche Diagnostics); Penn and Emory: Luminex xMAP INNO-BIA AlzBio3 (Luminex, Bio-techne); SPIN: Lumipulse G600, Fujirebio). Positive AD biomarker profile was determined by using predefined cut-offs (ADC: tTau/Aβ42 ratio > 0.46; Penn: tTau/Aβ42 ratio > 0.30; Emory: Aβ42/tTau < 6; SPIN: < 0.062 Aβ42/Aβ40 ratio, tTau > 456 pg/mL and pTau > 63 pg/mL). In the ADC, Innotest Aβ42 concentrations were adjusted for drift over time [83]. CSF biomarkers measured on Elecsys were transformed to Innotest values using conversion formulas using Passing-Bablok regression analysis based on cases from which both Luminex and Innotest values were available, as described previously [27]. CSF NfL was measured in a subset of cases from the ADC (discovery cohort: 42 CON, 74 FTD, and 54 AD; clinical cohort 1: 8 CON, 19 FTD, and 20 AD) either by NF-light® ELISA (Uman Diagnostics, Sweden) or with the single molecular array (Simoa®) NF-light™ advantage kit (Quanterix, USA)[84]. NfL concentrations measured by ELISA were converted to Simoa values using Passing-Bablok regression analysis, as described previously [84].

### Ethical statement

Approval was given by the institutional ethical review boards of each center. Written informed consent was obtained from all participants or their authorized representatives.

### CSF protein profiling

979 CSF proteins were quantified using 11 Olink Target 96 validated multiplex panels based on proximity extension assay (PEA) technology (Olink Proteomics, Uppsala, Sweden) that were available at the time of analysis (Cardiometabolic, Cardiovascular II and III, cell regulation, development, immune response, inflammation, metabolism, neurology, oncology II and organ damage). CSF samples were randomized across multiple plates containing intra- and inter-plate quality controls (QCs) from the manufacturer and measured in two different rounds. Each round of measurement contained 16 bridging samples covering different clinical groups, which were used as a reference to account for potential batch effects. For each protein, the lower limit of detection (LOD) was determined by the company, and defined as three standard deviations above the background from the negative controls included on every plate. Proteins were excluded from further analysis if levels were below the LOD in 15% of the samples. A total of 665 proteins (642 unique proteins) were ultimately included for statistical analysis of the discovery cohort, similar to our previous study [27]. Protein abundance was reported in normalized protein expression (NPX) values.

### Development of custom PEA assays

Multiplex-PEA assays were custom-developed by the manufacturer following standardized protocols [85]. We developed assays to measure 18 out of 24 proteins selected by the classification analysis described below. CSF samples from validation cohorts were randomized across plates. Each plate additionally included: four CSF QC samples, a negative control, and three calibrators used for normalization. Each custom assay had a LOD determined by the company, defined as three standard deviations above the background from the negative controls. Precision (intra- and inter-assay CV) was calculated using the four CSF QC samples (Supplementary Table 1). No cross-reactivity between assays for the specific proteins was detected. Samples from the validation cohorts were randomized across plates and normalized for any plate effects using the built-in inter-plate controls according to the manufacturer’s recommendations. Protein levels are reported in NPX values.

### Statistical analyses

All processing and statistical analysis were performed in R version 4.2.1. Baseline demographics were tested by Kruskal-Wallis test followed by Bonferonni post-hoc, or Peasons’s chi-square test for continuous and categorical variables, respectively. Differences in protein abundance of the CSF proteome data was tested with nested linear models including age and sex in the model for the comparison between different groups (FTD vs controls, FTD vs AD, FTLD-Tau vs controls, FTLD-TDP vs controls, and FTLD-Tau vs FTLD-TDP) as previously performed [27, 34]. For each pairwise comparison, multiplicity was taken into account by controlling the False Discovery Rate (FDR) at *q* ≤ 0.05 based on the number of features analyzed [86].

We next evaluated which CSF protein combination (CSF panels) could best discriminate the groups of interest while keeping the number of markers to a minimum so that they can be ultimately translated into small, practical custom panels, as described previously [27]. For this purpose, binary classification signatures (FTD vs. Controls, FTD vs. AD, and FTLD-Tau vs. FTLD-TDP) were constructed by way of penalized generalized linear modeling (GLM) with an elastic net penalty (a linear combination of lasso and ridge penalties) in the discovery cohort using the glmnet package, including age and sex as covariates in the model [27, 87, 88]. This penalty enables estimation in settings where the feature-to-sample ratio is too high for standard generalized linear regression. Moreover, it performs automatic feature decorrelation as well as feature-selection. For each classification exercise, we compare multiple models that reflect (a) a grid of values for the elastic-net mixing parameter, reflecting strong decorrelation to a pure logistic lasso regression, and (b) a grid of values reflecting the maximum number of proteins that may be selected under each model (21 markers maximum). The former grid (a) considers that we have little information on the collinearity burden in the data. The latter grid (b) considers that we want to keep the number of selected proteins relatively low for the future development of customized panels. The optimal penalty parameters in the penalized models were determined based on (balanced) 10-fold cross-validation of the model likelihood [27, 87]. The cross-validation was performed with balanced folds, by which each fold has an outcome group ratio close to the corresponding ratio in the full data set, also referred to as stratified cross-validation. The predictive performance of all models was assessed by way of (the comparison of) Receiver Operating Characteristic (ROC) curves and Area Under the ROC Curves (AUCs). The model with the highest AUC and lowest number of markers for each classification signature was selected. The fold-based selection proportions for each marker were assessed to identify and select the most promising markers within each model (i.e., features that are stably selected across each individual fold thereby minimizing potential overfitting). To reflect the manual selection pressure for these final marker sets, each final logistic signature was subjected to a ridge-regularization with a penalty parameter of 0.1. The performance (AUC) was evaluated by internal validation: repeated 5-fold cross-validation with 1000 repeats. The 95% confidence interval around the resulting AUCs was based on resampling quantiles (percentile method). External validations assessed the performance of the final models with the markers of interest in the validation cohorts. We additionally compared our identified classification models to CSF NfL to discriminate FTD from controls and AD in the subset of cases for which this information was available.

Functional enrichment analysis was performed for all comparisons described above using Metascape selecting GO biological Processes as an ontology source [89]. The total number of CSF proteins optimally analyzed was included as the enrichment background (n = 642). Default parameters were used for the analysis in which terms with a *p*-value < 0.01, a minimum count of 3, and an enrichment factor > 1.5 were collected and grouped into clusters based on their membership similarities.

Partial non-parametric correlation analysis was performed to understand the association among proteins within the CSF panels with cognitive function (MMSE score) and disease severity (FTLD-CDR® plus NACC scores)[81]. This analysis was corrected for age, sex, and the clinician who performed the FTLD-CDR examination as a dummy variable; and performed in the total discovery cohort and specifically for the FTD group.

## Data availability

The data generated in this study is available from the authors on reasonable request.

## Data Availability

The data generated in this study is available from the authors on reasonable request.

## Acknowledgements

This project was funded by the memorable project PRIDE (Project No. WE.03-2018-05), supported by Alzheimer Nederland. MdC is supported by the attraction talent fellowship from Comunidad de Madrid (2018-T2/BMD-11885). Data from Penn were contributed by the program project grants P01-AG-066597, U19 AG062418 (formerly P50 NS053488), and P30AG072979 (formerly AG010124).

## Author contributions

Y.H., M.C., and C.T. conceived and designed the study. Y.H, L.V., and C.P. performed the statistical analysis. Y.H., L.V., E.E., S.B., L.M., J.S., A.L., D.A., S.E., A.S., A.C.-P., D.I., W.F., Y.P., M.C., and C.T. recruited participants and collected clinical data and samples. Y.H. and M.C. arranged and prepared samples for proteomics analysis. Y.H., M.C., and C.T. drafted de manuscript. All authors contributed to the revision and editing of the manuscript.

## References

1. Gorno-Tempini, M.L., et al., Classification of primary progressive aphasia and its variants. American Academy of Neurology, 2011. 76.

2. Rascovsky, K., et al., Sensitivity of revised diagnostic criteria for the behavioural variant of frontotemporal dementia. Brain, 2011. 134(Pt 9): p. 2456–77.

3. Armstrong, M.J., et al., Criteria for the diagnosis of corticobasal degeneration. American Academy of Neurology, 2013. 80.

4. Hoglinger, G.U., et al., Clinical diagnosis of progressive supranuclear palsy: The movement disorder society criteria. Mov Disord, 2017. 32(6): p. 853–864.

5. Irwin, D.J., et al., Frontotemporal lobar degeneration: defining phenotypic diversity through personalized medicine. Acta Neuropathol, 2015. 129(4): p. 469–91.

6. Mann, D.M.A. and J.S. Snowden, Frontotemporal lobar degeneration: Pathogenesis, pathology and pathways to phenotype. Brain Pathol, 2017. 27(6): p. 723–736.

7. Neumann, M., et al., A new subtype of frontotemporal lobar degeneration with FUS pathology. Brain, 2009. 132(Pt 11): p. 2922–31.

8. Lashley, T., et al., Review: an update on clinical, genetic and pathological aspects of frontotemporal lobar degenerations. Neuropathol Appl Neurobiol, 2015. 41(7): p. 858–81.

9. Rohrer, J.D., et al., The heritability and genetics of frontotemporal lobar degeneration. Neurology, 2009. 73.

10. Greaves, C.V. and J.D. Rohrer, An update on genetic frontotemporal dementia. J Neurol, 2019. 266(8): p. 2075–2086.

11. Neumann, M., E.B. Lee, and I.R. Mackenzie, Frontotemporal Lobar Degeneration TDP-43-Immunoreactive Pathological Subtypes: Clinical and Mechanistic Significance. Adv Exp Med Biol, 2021. 1281: p. 201–217.

12. Del Campo, M., et al., New developments of biofluid-based biomarkers for routine diagnosis and disease trajectories in frontotemporal dementia. Alzheimers Dement, 2022. 18(11): p. 2292–2307.

13. Hutchinson, A.D. and J.L. Mathias, Neuropsychological deficits in frontotemporal dementia and Alzheimer’s disease: a meta-analytic review. J Neurol Neurosurg Psychiatry, 2007. 78(9): p. 917–28.

14. Naasan, G., et al., Amyloid in dementia associated with familial FTLD: not an innocent bystander. Neurocase, 2016. 22(1): p. 76–83.

15. Irwin, D.J., J.Q. Trojanowski, and M. Grossman, Cerebrospinal fluid biomarkers for differentiation of frontotemporal lobar degeneration from Alzheimer’s disease. Front Aging Neurosci, 2013. 5: p. 6.

16. Ashton, N.J., et al., A multicentre validation study of the diagnostic value of plasma neurofilament light. Nat Commun, 2021. 12(1): p. 3400.

17. Bridel, C., et al., Diagnostic Value of Cerebrospinal Fluid Neurofilament Light Protein in Neurology: A Systematic Review and Meta-analysis. JAMA Neurol, 2019. 76(9): p. 1035–1048.

18. Ducharme, S., et al., Recommendations to distinguish behavioural variant frontotemporal dementia from psychiatric disorders. Brain, 2020. 143(6): p. 1632–1650.

19. Gendron, T.F., et al., Comprehensive cross-sectional and longitudinal analyses of plasma neurofilament light across FTD spectrum disorders. Cell Rep Med, 2022. 3(4): p. 100607.

20. Meeter, L.H.H., et al., Clinical value of neurofilament and phospho-tau/tau ratio in the frontotemporal dementia spectrum. Neurology, 2018. 90(14): p. e1231–e1239.

21. Pijnenburg, Y.A., et al., Discriminative and prognostic potential of cerebrospinal fluid phosphoTau/tau ratio and neurofilaments for frontotemporal dementia subtypes. Alzheimers Dement (Amst), 2015. 1(4): p. 505–12.

22. Cousins, K.A.Q., et al., Distinguishing Frontotemporal Lobar Degeneration Tau From TDP-43 Using Plasma Biomarkers. JAMA Neurol, 2022. 79(11): p. 1155–1164.

23. Desmarais, P., et al., Therapeutic trial design for frontotemporal dementia and related disorders. J Neurol Neurosurg Psychiatry, 2019. 90(4): p. 412–423.

24. Higginbotham, L., et al., Integrated proteomics reveals brain-based cerebrospinal fluid biomarkers in asymptomatic and symptomatic Alzheimer’s disease. Science Advances, 2020. 6(43).

25. Bader, J.M., et al., Proteome profiling in cerebrospinal fluid reveals novel biomarkers of Alzheimer’s disease. Mol Syst Biol, 2020. 16(6): p. e9356.

26. Dammer, E.B., et al., Multi-platform proteomic analysis of Alzheimer’s disease cerebrospinal fluid and plasma reveals network biomarkers associated with proteostasis and the matrisome. Alzheimers Res Ther, 2022. 14(1): p. 174.

27. Del Campo, M., et al., CSF proteome profiling across the Alzheimer’s disease spectrum reflects the multifactorial nature of the disease and identifies specific biomarker panels. Nature Aging, 2022. 2(11): p. 1040–1053.

28. Teunissen, C.E., et al., Novel diagnostic cerebrospinal fluid biomarkers for pathologic subtypes of frontotemporal dementia identified by proteomics. Alzheimers Dement (Amst), 2016. 2: p. 86–94.

29. van der Ende, E.L., et al., Novel CSF biomarkers in genetic frontotemporal dementia identified by proteomics. Ann Clin Transl Neurol, 2019. 6(4): p. 698–707.

30. Del Campo, M., et al., Novel CSF biomarkers to discriminate FTLD and its pathological subtypes. Ann Clin Transl Neurol, 2018. 5(10): p. 1163–1175.

31. van der Ende, E.L., et al., Neuronal pentraxin 2: a synapse-derived CSF biomarker in genetic frontotemporal dementia. J Neurol Neurosurg Psychiatry, 2020. 91(6): p. 612–621.

32. Bolsewig, K., et al., A Combination of Neurofilament Light, Glial Fibrillary Acidic Protein, and Neuronal Pentraxin-2 Discriminates Between Frontotemporal Dementia and Other Dementias. J Alzheimers Dis, 2022. **90**(1): p. 363-380.

33. Llorens, F., et al., YKL-40 in the brain and cerebrospinal fluid of neurodegenerative dementias. Mol Neurodegener, 2017. 12(1): p. 83.

34. Del Campo, M., et al., CSF proteome profiling reveals biomarkers to discriminate dementia with Lewy bodies from Alzheimer s disease. Nat Commun, 2023. 14(1): p. 5635.

35. Huang, X., et al., VSIG4 mediates transcriptional inhibition of Nlrp3 and Il-1b in macrophages. Science Advances, 2019. 5.

36. Zhang, J.Q., et al., Siglec-9, a novel sialic acid binding member of the immunoglobulin superfamily expressed broadly on human blood leukocytes. J Biol Chem, 2000. 275(29): p. 22121–6.

37. Yan, Q., et al., Structure of CD84 provides insight into SLAM family function. PNAS, 2007. 104(25).

38. Tanaka, Y., et al., Progranulin regulates lysosomal function and biogenesis through acidification of lysosomes. Hum Mol Genet, 2017. 26(5): p. 969–988.

39. Kishore, U., et al., C1q and tumor necrosis factor superfamily: modularity and versatility. Trends Immunol, 2004. 25(10): p. 551–61.

40. Goel, M., et al., Cochlin induced TREK-1 co-expression and annexin A2 secretion: role in trabecular meshwork cell elongation and motility. PLoS One, 2011. 6(8): p. e23070.

41. Karlsson, L., et al., Cerebrospinal fluid reference proteins increase accuracy and interpretability of biomarkers for brain diseases. bioRxiv, 2023.

42. Poe, M., et al., Human cytotoxic lymphocyte granzyme B. Its purification from granules and the characterization of substrate and inhibitor specificity. Journal of Biological Chemistry, 1991. 266(1): p. 98–103.

43. Mroczko, B., M. Groblewska, and M. Barcikowska, The role of matrix metalloproteinases and tissue inhibitors of metalloproteinases in the pathophysiology of neurodegeneration: a literature study. J Alzheimers Dis, 2013. 37(2): p. 273–83.

44. Spencer, M.L., M. Theodosiou, and D.J. Noonan, NPDC-1, a novel regulator of neuronal proliferation, is degraded by the ubiquitin/proteasome system through a PEST degradation motif. J Biol Chem, 2004. 279(35): p. 37069–78.

45. Pei, D.S., et al., AP endonuclease 1 (Apex1) influences brain development linking oxidative stress and DNA repair. Cell Death Dis, 2019. 10(5): p. 348.

46. Olsson, A.K., et al., VEGF receptor signalling - in control of vascular function. Nat Rev Mol Cell Biol, 2006. 7(5): p. 359–71.

47. Bostrom, G., et al., Different Inflammatory Signatures in Alzheimer’s Disease and Frontotemporal Dementia Cerebrospinal Fluid. J Alzheimers Dis, 2021. 81(2): p. 629–640.

48. Mol, M.O., et al., Proteomics of the dentate gyrus reveals semantic dementia specific molecular pathology. Acta Neuropathol Commun, 2022. 10(1): p. 190.

49. Miedema, S.S.M., et al., Distinct cell type-specific protein signatures in GRN and MAPT genetic subtypes of frontotemporal dementia. Acta Neuropathol Commun, 2022. 10(1): p. 100.

50. Umoh, M.E., et al., A proteomic network approach across the ALS-FTD disease spectrum resolves clinical phenotypes and genetic vulnerability in human brain. EMBO Mol Med, 2018. 10(1): p. 48–62.

51. Mofrad, R.B., et al., Plasma proteome profiling identifies changes associated to AD but not to FTD. Acta Neuropathol Commun, 2022. 10(1): p. 148.

52. Hok-A-Hin, Y.S., et al., Neuroinflammatory CSF biomarkers MIF, sTREM1, and sTREM2 show dynamic expression profiles in Alzheimer’s disease. J Neuroinflammation, 2023. **20**(1): p. 107.

53. Zhang, S., et al., Upregulation of MIF as a defense mechanism and a biomarker of Alzheimer’s disease. Alzheimers Res Ther, 2019. 11(1): p. 54.

54. Remnestal, J., et al., Altered levels of CSF proteins in patients with FTD, presymptomatic mutation carriers and non-carriers. Transl Neurodegener, 2020. 9(1): p. 27.

55. Wise, A., et al., CSF Proteomics in Patients With Progressive Supranuclear Palsy. Neurology, 2024. 103(3): p. e209585.

56. Martino Adami, P.V., et al., Matrix metalloproteinase 10 is linked to the risk of progression to dementia of the Alzheimer’s type. Brain, 2022. 145(7): p. 2507–2517.

57. Abu-Rumeileh, S., et al., CSF biomarkers of neuroinflammation in distinct forms and subtypes of neurodegenerative dementia. Alzheimers Res Ther, 2019. 12(1): p. 2.

58. Steinacker, P., et al., Chitotriosidase (CHIT1) is increased in microglia and macrophages in spinal cord of amyotrophic lateral sclerosis and cerebrospinal fluid levels correlate with disease severity and progression. J Neurol Neurosurg Psychiatry, 2018. 89(3): p. 239–247.

59. van der Ende, E.L., et al., CSF proteomics in autosomal dominant Alzheimer’s disease highlights parallels with sporadic disease. Brain, 2023.

60. Whelan, C.D., et al., Multiplex proteomics identifies novel CSF and plasma biomarkers of early Alzheimer’s disease. Acta Neuropathol Commun, 2019. 7(1): p. 169.

61. Guo, Y., et al., Multiplex cerebrospinal fluid proteomics identifies biomarkers for diagnosis and prediction of Alzheimer’s disease. Nat Hum Behav, 2024.

62. Bovolenta, P., et al., Beyond Wnt inhibition: new functions of secreted Frizzled-related proteins in development and disease. J Cell Sci, 2008. 121(Pt 6): p. 737–46.

63. Hadi, F., et al., Wnt signalling pathway and tau phosphorylation: A comprehensive study on known connections. Cell Biochem Funct, 2020. 38(6): p. 686–694.

64. Sogorb-Esteve, A., et al., Differential impairment of cerebrospinal fluid synaptic biomarkers in the genetic forms of frontotemporal dementia. Alzheimers Res Ther, 2022. 14(1): p. 118.

65. 65. Saloner, R., *Large-scale network analysis of the cerebrospinal fluid proteome identifies molecular signatures of frontotemporal lobar degeneration.* Preprint Biorxv, 2024.

66. Goossens, J., et al., Diagnostic value of cerebrospinal fluid tau, neurofilament, and progranulin in definite frontotemporal lobar degeneration. Alzheimers Res Ther, 2018. 10(1): p. 31.

67. Meda, F., et al., Analytical and clinical validation of a blood progranulin ELISA in frontotemporal dementias. Clin Chem Lab Med, 2023.

68. Davis, S.E., et al., Patients with sporadic FTLD exhibit similar increases in lysosomal proteins and storage material as patients with FTD due to GRN mutations. Acta Neuropathol Commun, 2023. 11(1): p. 70.

69. Mackenzie, I.R., et al., The neuropathology of frontotemporal lobar degeneration caused by mutations in the progranulin gene. Brain, 2006. 129(Pt 11): p. 3081–90.

70. Ising, C., et al., NLRP3 inflammasome activation drives tau pathology. Nature, 2019. 575(7784): p. 669-673.

71. Palluzzi, F., et al., A novel network analysis approach reveals DNA damage, oxidative stress and calcium/cAMP homeostasis-associated biomarkers in frontotemporal dementia. PLoS One, 2017. 12(10): p. e0185797.

72. Malpetti, M., et al., Microglial activation in the frontal cortex predicts cognitive decline in frontotemporal dementia. Brain, 2023. 146(8): p. 3221–3231.

73. Nuttall, R.K., et al., Metalloproteinases are enriched in microglia compared with leukocytes and they regulate cytokine levels in activated microglia. Glia, 2007. 55(5): p. 516–26.

74. Lleo, A., et al., A 2-Step Cerebrospinal Algorithm for the Selection of Frontotemporal Lobar Degeneration Subtypes. JAMA Neurol, 2018. 75(6): p. 738–745.

75. van der Flier, W.M. and P. Scheltens, Amsterdam Dementia Cohort: Performing Research to Optimize Care. J Alzheimers Dis, 2018. 62(3): p. 1091–1111.

76. Ingelsson, M., et al., Increase in the relative expression of tau with four microtubule binding repeat regions in frontotemporal lobar degeneration and progressive supranuclear palsy brains. Acta Neuropathol, 2007. 114(5): p. 471–9.

77. Alcolea, D., et al., The Sant Pau Initiative on Neurodegeneration (SPIN) cohort: A data set for biomarker discovery and validation in neurodegenerative disorders. Alzheimers Dement (N Y), 2019. 5: p. 597–609.

78. Dubois, B., et al., Research criteria for the diagnosis of Alzheimer’s disease: revising the NINCDS-ADRDA criteria. Lancet Neurol, 2007. 6(8): p. 734–46.

79. Cairns, N.J., et al., Neuropathologic diagnostic and nosologic criteria for frontotemporal lobar degeneration: consensus of the Consortium for Frontotemporal Lobar Degeneration. Acta Neuropathol, 2007. 114(1): p. 5–22.

80. Montine, T.J., et al., National Institute on Aging-Alzheimer’s Association guidelines for the neuropathologic assessment of Alzheimer’s disease: a practical approach. Acta Neuropathol, 2012. 123(1): p. 1–11.

81. Miyagawa, T., et al., Utility of the global CDR plus NACC FTLD rating and development of scoring rules: Data from the ARTFL/LEFFTDS Consortium. Alzheimers Dement, 2020. 16(1): p. 106–117.

82. Hok-A-Hin, Y.S., et al., Guidelines for CSF Processing and Biobanking: Impact on the Identification and Development of Optimal CSF Protein Biomarkers. Methods Mol Biol, 2019. 2044: p. 27–50.

83. Tijms, B.M., et al., Unbiased Approach to Counteract Upward Drift in Cerebrospinal Fluid Amyloid-beta 1-42 Analysis Results. Clin Chem, 2018. 64(3): p. 576–585.

84. Vermunt, L., et al., Age- and disease-specific reference values for neurofilament light presented in an online interactive support interface. Ann Clin Transl Neurol, 2022. 9(11): p. 1832–1837.

85. Proteomics, O., Development and validation of customized PEA biomarker panels with clinical utility.

86. Benjamini, Y. and Y. Hochberg, Controlling the False Discovery Rate: A Practical and Powerful Approach to Multiple Testing. Journal of the Royal Statistical Society. Series B (Methodological), 1995. 57.

87. de Leeuw, F.A., et al., Blood-based metabolic signatures in Alzheimer’s disease. Alzheimers Dement (Amst), 2017. 8: p. 196–207.

88. Friedman, J., T. Hastie, and R. Tibshirani, Regularization Paths for Generalized Linear Models via Coordinate Descent. J Stat Softw, 2010. 33(1): p. 22.

89. Zhou, Y., et al., Metascape provides a biologist-oriented resource for the analysis of systems-level datasets. Nat Commun, 2019. 10(1): p. 1523.

