## Supplemental File for "Large-scale CSF proteome profiling identifies biomarkers for accurate diagnosis of Frontotemporal Dementia"

**Supplemental File 1.**

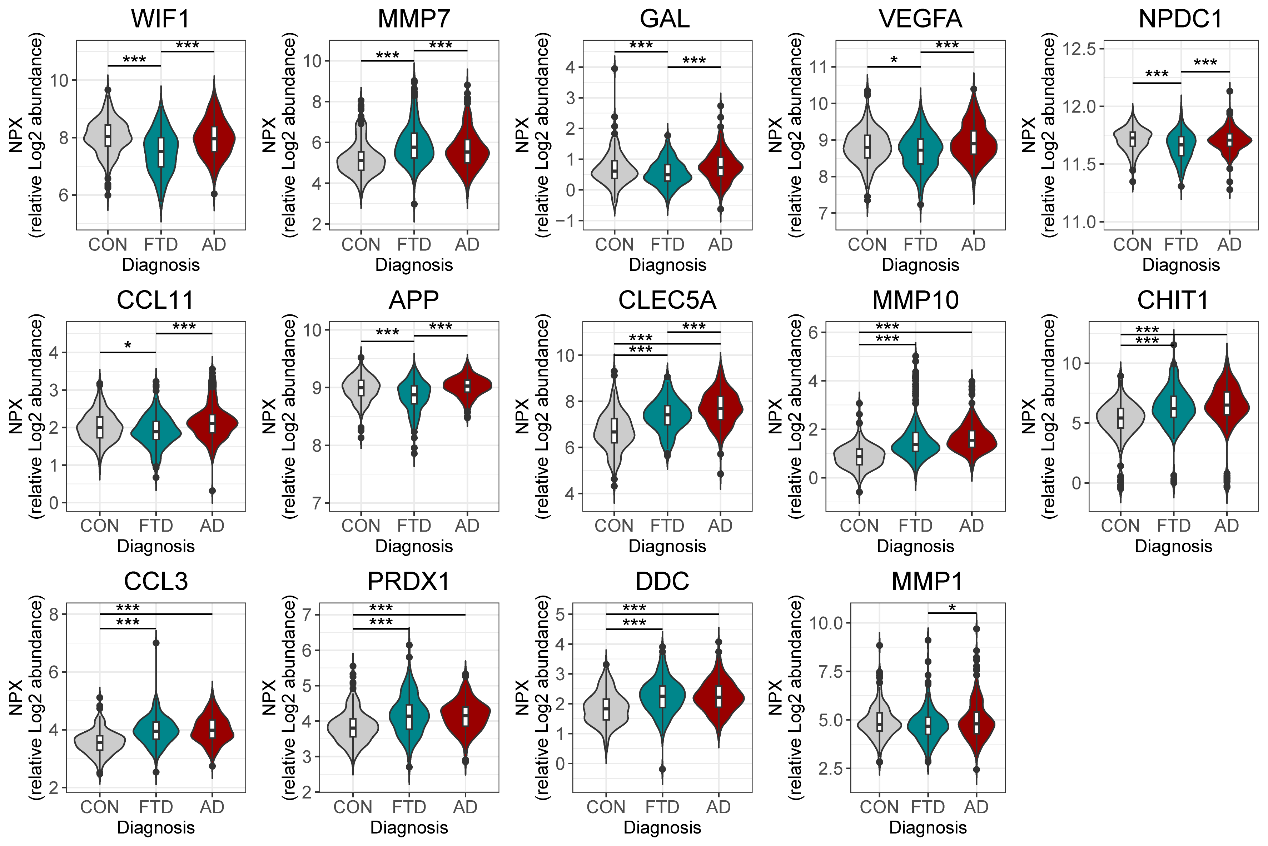

Supplementary Figure 1. Protein levels of FTD diagnostic biomarker panel.

Violins represent the abundance (log2 NPX) of the CSF proteins that combined can accurately discriminate FTD from AD. Boxplot within the violin indicates the median and interquartile range of the protein abundance.*q < 0.05, **q < 0.01, ***q < 0.001. n.s: non-significant. Abbreviations: CON, cognitively unimpaired controls; FTD, frontotemporal dementia; AD, Alzheimer’s disease

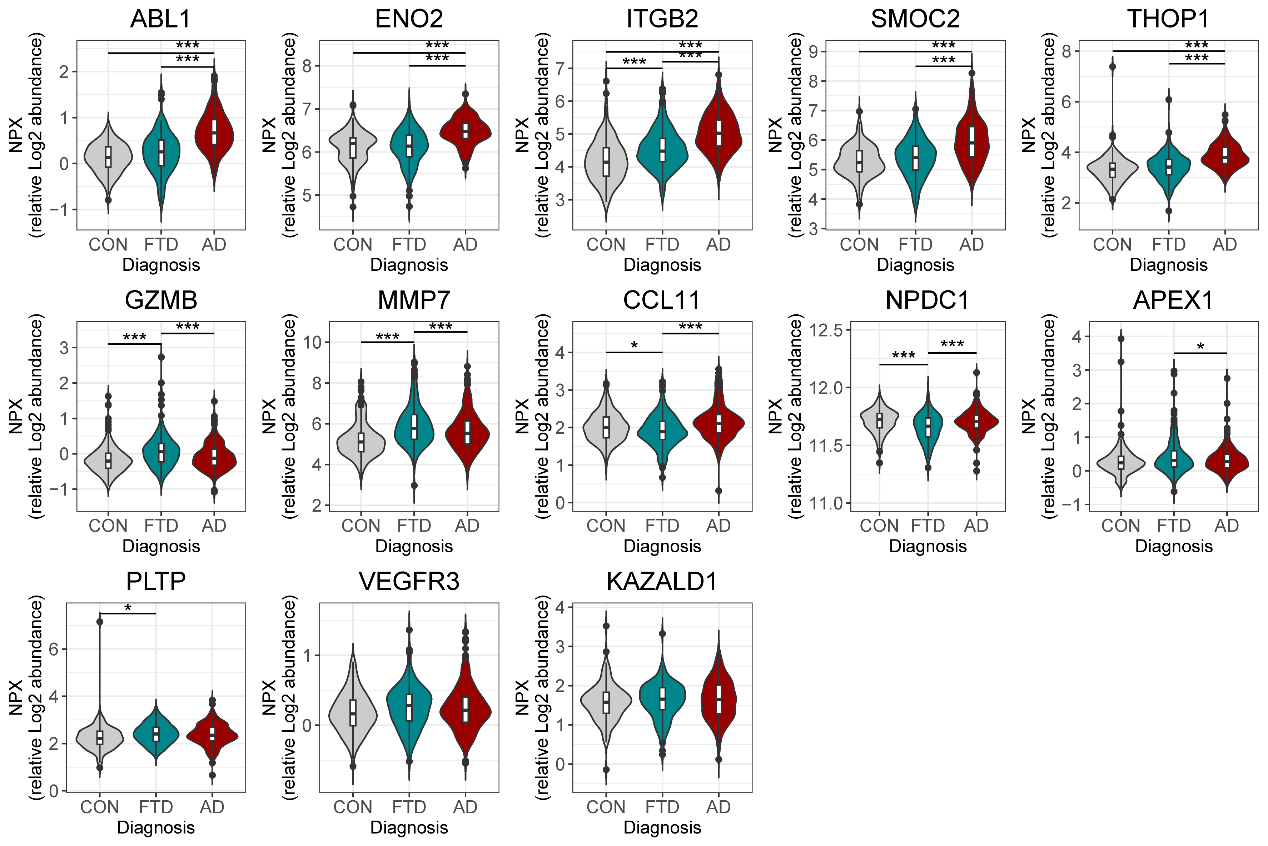

Supplementary Figure 2. Protein levels of FTD differential diagnostic biomarker panel.

Violins represent the abundance (log2 NPX) of the CSF proteins that combined can accurately discriminate FTD from controls. Boxplot within the violin indicates the median and interquartile range of the protein abundance.*q < 0.05, **q < 0.01, ***q < 0.001. n.s: non-significant. Abbreviations: CON, cognitively unimpaired controls; FTD, frontotemporal dementia; AD, Alzheimer’s disease

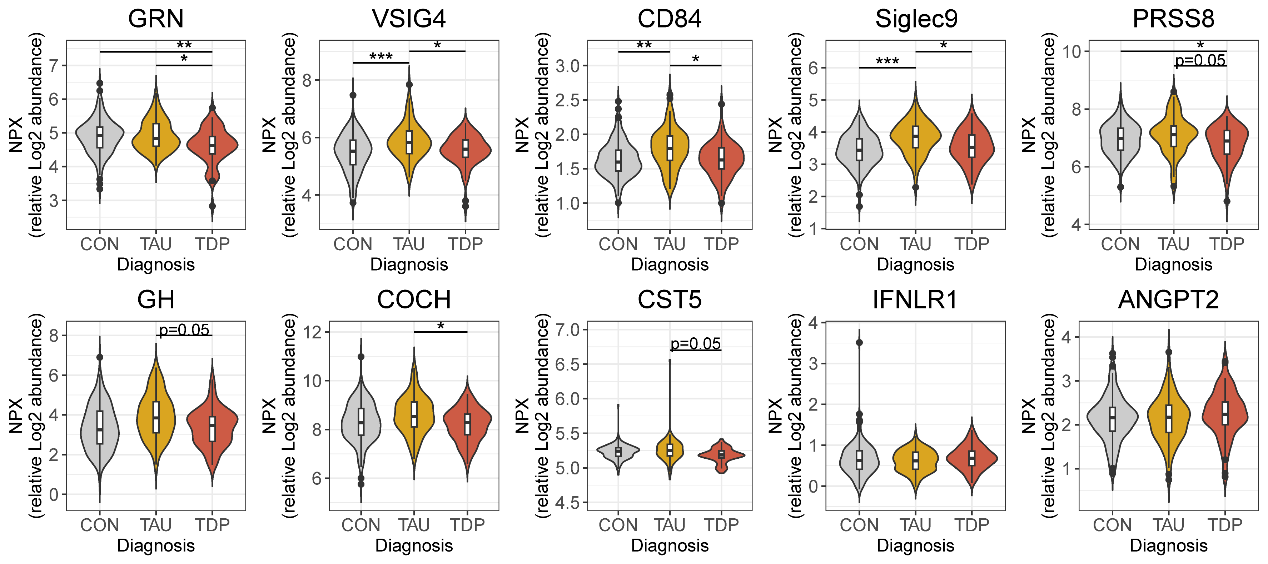

Supplementary Figure 3. Protein levels of FTLD subtype biomarker panel.

Violins represent the abundance (log2 NPX) of the CSF proteins that combined can accurately discriminate FTLD-Tau from FTLD-TDP. Boxplot within the violin indicates the median and interquartile range of the protein abundance.*q < 0.05, **q < 0.01, ***q < 0.001. n.s: non-significant. Abbreviations: CON, cognitively unimpaired controls; TDP, Transactive response DNA binding protein of 43.

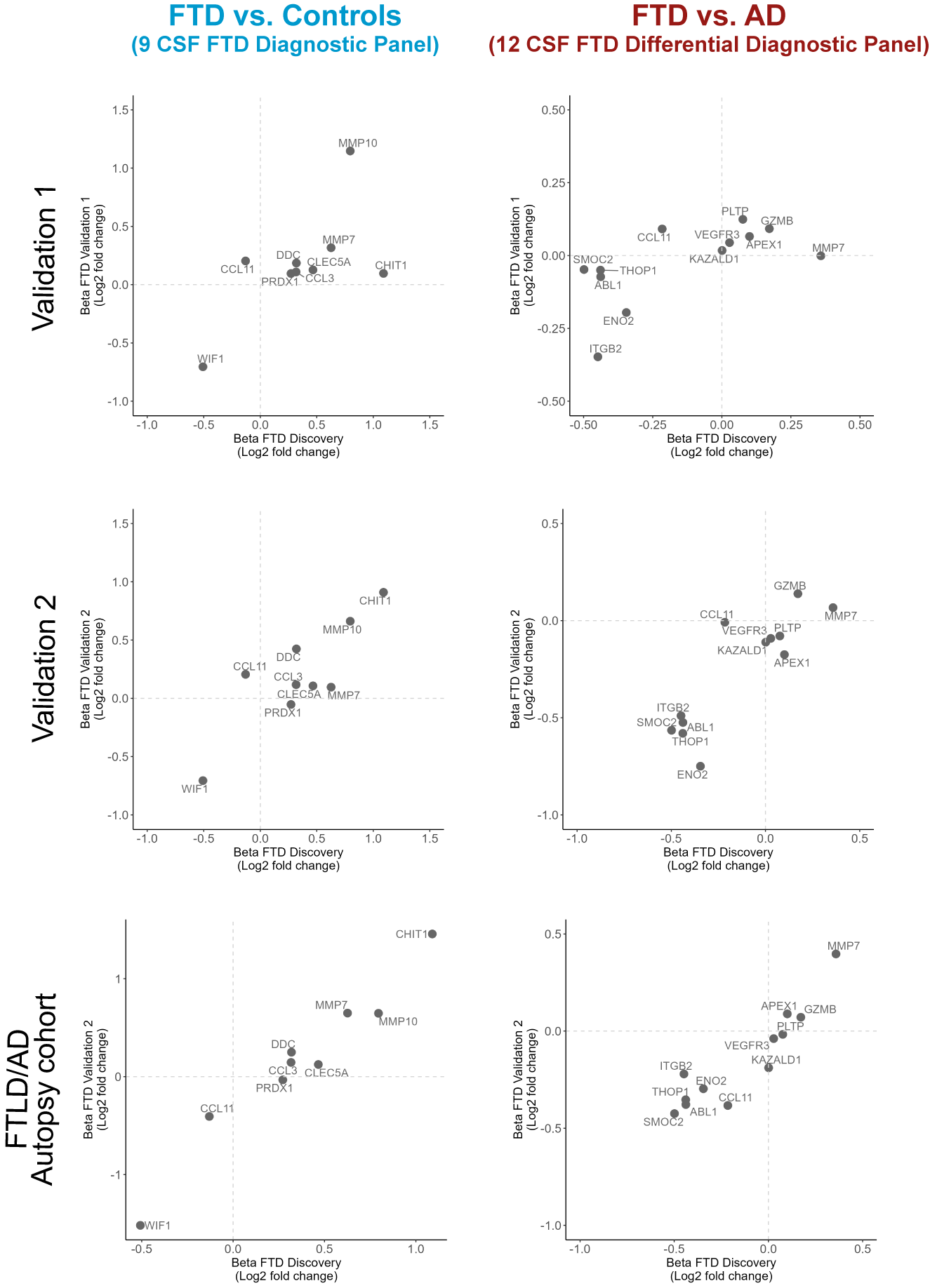

Supplementary Figure 4. Correlations between discovery and validation cohorts.

Scatter plots depict the correlation between the beta-coefficients obtained in the discovery phase to those obtained with the custom assays in the clinical validation cohorts 1 and 2 and the FTLD/AD autopsy cohort.
